# Altered oscillatory coupling and a candidate inhibitory circuit mechanism in Rett syndrome

**DOI:** 10.1101/2025.07.21.25331927

**Authors:** Devorah Kranz, Yael Braverman, Michelle McCarthy, Karen N Sabol, Tim A. Benke, David N Lieberman, Eric D. Marsh, Jeffrey L Neul, Fleming Peck, Alan K Percy, Joni Saby, Damla Senturk, Nancy Kopell, Charles A. Nelson, April R Levin, Michela Fagiolini

## Abstract

Rett syndrome (RTT) is a rare neurodevelopmental disorder caused primarily by pathogenic variants in the MECP2 gene and characterized by severe cognitive and neurological impairment. Phase–amplitude coupling (PAC), a cross-frequency interaction supporting information integration across temporal and spatial scales, provides a window into large-scale neural coordination and circuit dysfunction. PAC depends on coordinated activity among specific neuronal subtypes, yet the cellular mechanisms shaping oscillatory dynamics in RTT remain poorly understood. Here we combined electroencephalography (EEG) with biophysically constrained computational modeling to characterize PAC alterations and investigate their circuit-level origins. Awake resting-state EEG was recorded from 42 individuals with RTT and 37 typically developing participants, and PAC strength was quantified. Individuals with RTT exhibited stronger PAC across widespread frequency pairs and cortical regions, including elevated theta–gamma coupling in posterior, midline, and whole-brain regions, and elevated alpha–gamma coupling in anterior, posterior, and whole-brain regions. Biophysically constrained network simulations of a cortical Layer 4 model comprising pyramidal neurons together with PV+, SOM+, and VIP+ interneurons demonstrated that reduced excitability of VIP+ interneurons selectively reproduced the increased theta–gamma and alpha–gamma PAC observed in RTT. These findings identify abnormal PAC as an electrophysiological feature of RTT and suggest a possible mechanistic link between altered oscillatory coordination and VIP+ interneuron dysfunction. PAC may provide a non-invasive marker of inhibitory circuit dysfunction and a translational framework for investigating shared circuit abnormalities across neurodevelopmental disorders.

**Significance Statement:** Rett syndrome has a known genetic cause, but the circuit changes linking cellular dysfunction to altered brain activity remain unclear. Combining electroencephalography with computational modeling, we found stronger coupling between slow and fast brain rhythms in Rett syndrome compared to typically developing controls and identified reduced excitability of VIP-expressing inhibitory neurons as one plausible mechanism. These findings connect a non-invasive measure of human brain activity with a candidate cellular mechanism. Beyond Rett syndrome, this approach could help identify subgroups with shared circuit dysfunction across neurodevelopmental disorders, supporting biologically informed participant stratification and the development of targeted interventions.

## Introduction

Rett syndrome (RTT) is a rare X-linked neurodevelopmental disorder that presents with an initial period of typical development followed by rapid regression resulting in profound cognitive, motor, and autonomic impairments including loss of purposeful hand use and spoken language (1, 2). The majority of RTT cases are caused by de novo pathogenic variants in the X-linked *MECP2* gene, which encodes methyl-CpG-binding protein 2 (MeCP2) (3). Although MeCP2 is expressed widely throughout the body, its levels are 5 to 10-fold higher in neurons compared to other cell types, and neuronal dysfunction is central to RTT pathology (4). MeCP2 influences the brain’s cellular and network development across the lifespan: its function affects prenatal neurogenesis, development of neuronal circuitry, and maintenance of adult neural function (5–8). Identifying scalable neurophysiological markers that reflect underlying circuit dysfunction may therefore provide a critical bridge between molecular pathology and systems-level brain abnormalities in RTT and related neurodevelopmental disorders (1, 9).

Brain-wide dynamics rely heavily on coordination of and by neural oscillations—rhythmic patterns of neuronal activity characterized by frequency, amplitude, and phase—that facilitate communication between distant cortical regions and support cognitive processing (10). Electroencephalography (EEG) provides a valuable non-invasive and clinically-relevant tool to assess these oscillations and is particularly suitable for use in RTT as it can be recorded without requiring active task engagement. Clinical EEG in RTT has long revealed visually observed abnormalities such as epileptiform discharges and diffuse slowing of activity (11–13). Recent quantitative studies have expanded upon these findings, revealing evidence of region-specific disturbances to spectral power (which examines the power of each individual frequency present in the signal), including increases in delta (1-3 Hz) and theta power (4-7 Hz), reductions in alpha power (8-12 Hz), and alterations in the 1/f spectral slope (14, 15). While these analyses offer insight into the overall magnitude of neural oscillations, they provide limited information about how oscillations dynamically interact to support cognitive processes, or how circuit-level dysfunction in RTT can disrupt communication between brain regions or alter precise timing mechanisms essential for cognition.

Phase-amplitude coupling (PAC), in which the phase of low-frequency oscillations modulates the amplitude of high-frequency activity, has emerged as a fundamental mechanism for coordinating neural communication across spatial and temporal scales (16–18). PAC has been observed in diverse electrophysiological signals, from intracellular recordings to scalp EEG, highlighting its relevance across different levels of neural activity. It is thought that PAC serves as a clocking mechanism, generating temporal windows that allow for integration of relevant information (19). Notably, PAC can vary by cortical layer and is influenced by interneuron function (20–22).

PAC between predefined frequency pairs is particularly relevant to specific domains of brain function and has been shown to be functionally important to a range of cognitive processes. Theta-gamma coupling is implicated in working memory, with theta phase modulating gamma amplitude to organize memory encoding and retrieval in the hippocampus and prefrontal cortex (23–26). Alpha-gamma coupling, meanwhile, has been linked to computations in visual cortical areas, where alpha phase modulates local gamma activity, facilitating selective attention, visual information integration, and prioritization of sensory input based on stimulus saliency (27). Altered PAC has been reported in autism spectrum disorder (ASD) and other neurogenetic disorders, both in resting-state EEG (28) and during cognitive tasks (29–33). In RTT, abnormal PAC has recently been identified during slow-wave sleep (34), but its presence during the awake state—particularly at faster frequencies associated with memory and attention—has not yet been studied. Moreover, precisely which neuronal subpopulations and circuit mechanisms drive RTT-related oscillatory abnormalities remains an area of active investigation.

In *Mecp2* mutant mice, which replicate key aspects of the RTT phenotype, cortical circuit dysfunction is characterized by an imbalance in excitatory-inhibitory signaling (35–37), and studies employing techniques such as selective deletion of *Mecp2* from neuronal subtypes have clarified how distinct cell populations individually contribute to these network disruptions (38–40). Pyramidal (PYR) neuron activity is significantly reduced, particularly in sensory cortical regions, leading to progressive silencing of visual circuits and decreased visual acuity (35, 37, 41). Alterations in GABAergic interneurons have also been strongly implicated (42): parvalbumin-positive interneurons (PV+INs), which normally provide fast-spiking inhibition to PYR neurons, exhibit increased connectivity and activity in *Mecp2* mutant mice (36). Vasoactive intestinal peptide-expressing interneurons (VIP+INs) have emerged as especially relevant to circuit and oscillatory dysfunction. VIP+INs synapse onto both PYR cells and other interneurons (43, 44), modulating sensory gain (45–49) and driving oscillatory activity (50, 51). Early disruptions in VIP+IN activity broadly compromise cortical circuits, resulting in weakened synaptic inhibition, impaired experience-dependent plasticity, and abnormal cortical maturation (45). Deficits in VIP+IN excitability and modulation have been reported in mouse models of other neurodevelopmental disorders (52, 53) that share some clinical features with RTT (54, 55). Notably, recent studies demonstrate that selective deletion of *Mecp2* from VIP+INs alters their intrinsic activity and recapitulates core phenotypic features of RTT (40), positioning VIP+IN dysfunction as a compelling avenue for understanding altered cortical dynamics in RTT.

In the present study, we adopted a multi-level approach, characterizing neural dynamics from both top-down and bottom-up perspectives. We first investigated phase-amplitude coupling patterns during awake resting state EEG in individuals with RTT. Since the specific frequencies and cortical regions where PAC may be disrupted in RTT are unknown, we initially conducted an exploratory, data-driven analysis examining coupling comprehensively across all sampled frequencies and scalp regions. Subsequently, we focused our analyses on two predefined frequency pairs—theta-gamma and alpha-gamma—given their established roles in cognitive processing. We identified abnormal PAC in individuals with RTT compared to typically developing controls, revealing significantly elevated coupling across multiple frequency bands and cortical regions, particularly in the alpha-gamma and theta-gamma ranges.

To identify potential neuronal and network mechanisms underlying the increased theta-gamma and alpha-gamma coupling seen in RTT, we then took a bottom-up approach by constructing a biophysically-constrained network model of cortical Layer 4 (L4) comprising pyramidal neurons and three key inhibitory interneuron classes: somatostatin (SOM+), PV+, and VIP+ interneurons. We focused our investigation on L4 for three reasons: (1) a major source of cortical alpha is thought to arise via thalamocortical interactions which include L4 (20, 56); (2) increased theta-gamma coupling has been associated with thalamocortical dysfunction (57); (3) interneurons are key mediators of oscillatory activity (58–60) and L4 consists of a relatively large 7/1 ratio of inhibitory to excitatory neuronal subtypes (61). Of note, biophysically-constrained models have been instrumental in revealing the connection of interneuron activity to rhythmic network dynamics (62–64). Here, our computational model suggests that decreased excitability of VIP+ interneurons alone could account for the observed PAC alterations in L4 dynamics.

By integrating clinical EEG findings with mechanistic insights from animal models and computational modeling, our approach provides a unique translational framework for linking electrophysiological measures with underlying cellular and circuit-level disruptions in RTT. Establishing PAC as a potential marker of inhibitory circuit dysfunction could enable more precise tracking of disease progression and therapeutic response in RTT and may provide a broadly applicable framework for linking cell-type–specific pathology to large-scale network dysfunction in neurodevelopmental disorders.

## Materials and Methods

### Participants

Participants were enrolled in one of two studies whose data were combined for the analyses presented here:

1. Natural History Study of Rett and Related Disorders (NHS) (U54 HD061222; NCT02738281) (65). Data were acquired across three study locations (Boston Children’s Hospital (BCH), Vanderbilt University Medical Center (VUMC), or Children’s Hospital Colorado (CHCO). The experimental protocol was approved and overseen by the Institutional Review Boards at each site.
2. Rett Syndrome Biomarkers Study (RSBS). Data were acquired only at BCH. The experimental protocol was approved and overseen by the Institutional Review Board at Boston Children’s Hospital.

Participants were 1-23-year-old individuals with Rett Syndrome (RTT; *n* = 53) and typical development (TD; *n* = 39). While many prior studies in RTT have focused on a narrower age range, we chose this wider range to capture the full spectrum of the disorder, and examined the effects of age on our analyses (described below). Participants were recruited through hospital-based clinics, a research registry, clinical referrals, community resources, and word of mouth. Informed consent was obtained from all guardians or participants above 18 years old, and assent from all participants developmentally able.

In the RTT group, inclusion criteria included being 12 months to 23 years old at time of recording and a diagnosis of RTT with an identified or likely pathogenic variant of *MECP2*. TD participants were approximately age and sex-matched and were included if they had no underlying genetic diagnoses or prior medical conditions associated with increased likelihood of ASD or intellectual disability. In both groups, the primary spoken language was English. Exclusion criteria for the RTT group included taking an investigational drug as part of another research study within 30 days prior to enrollment.

Participant characteristics are reported in Table 1.

**Table 1.** Participant information.

|  | **Diagnostic Group** |  |
|---|---|---|
|  | **RTT (n = 42)** | **TD (n=37)** |
| **Clinical Severity Score** |  |  |
| <10 | 3 |  |
| 10-19 | 16 |  |
| 20-29 | 19 |  |
| 30-39 | 4 |  |
| **Rett Diagnosis** |  |  |
| Typical | 33 |  |
| Atypical | 9 |  |
| **Seizure Status** |  |  |
| None | 23 |  |
| None (controlled with medication) | 10 |  |
| Monthly | 5 |  |
| Weekly | 3 |  |
| Daily | 1 |  |
| **MeCP2 Mutation** |  |  |
| Yes | 42 |  |
| No | 0 |  |
| **Sex** |  |  |
| Female | 39 | 35 |
| Male | 3 | 2 |
| **Age** | Range: 1-23 | Range: 1-23 |
|  | Median: 7 | Median: 7 |
|  | Mean: 9.2 (SD 6.8) | Mean: 8.3 (SD 5.9) |
| **Collection Site** |  |  |
| Vanderbilt | 11 | 1 |
| BCH | 30 | 32 |
| Denver | 1 | 4 |
| **Study** |  |  |
| RSBS | 27 | 26 |
| NHS | 15 | 11 |

### EEG data collection

Participants watched an approximately five-minute video directing their attention to the monitor, consisting of either repeating silent videos akin to screensavers or a silent video of their choice. A behavioral assistant sat in the room with the participant to keep them calm. Continuous EEG data were recorded at “rest” (with no time-locked stimuli) during the awake quiet state in dim lighting via a Net Amps 400 amplifier using the 128-channel Hydrocel Geodesic Net (Electrical Geodesic, Inc., Eugene, OR) with a sampling rate of 1000 Hz.

### EEG data pre-processing

EEG data preprocessing was conducted using the Batch EEG Automated Processing Platform (BEAPP) (66). Within BEAPP, data were artifact detected and corrected using the Harvard Automated Processing Pipeline for Electroencephalography (HAPPE) 1.0, a pipeline optimized for EEG data collected from children with neurodevelopmental disorders (67). Data were first bandpass filtered using a high pass (1 Hz) and low pass (100 Hz) filter and resampled to 250 Hz. Only the 18 electrodes in the international 10–20 system were chosen for further analysis. HAPPE applied line noise removal at 60 Hz via CleanLine’s multi-taper method and then automatically detected and rejected bad channels, movement, muscle, and eye blink artifact with wavelet-enhanced independent component analysis (ICA) and ICA with Multiple Artifact Rejection Algorithm (MARA) (68, 69). Following artifact removal, bad channels were interpolated using spherical interpolation and data were re-referenced to an average reference. Data were then segmented into overlapping 6 second segments (20% overlap), and a 2-second buffer was included at the beginning and end of each EEG segment to avoid edge effects from filters (the buffer resulted in 10-second segments, but only the middle 6 seconds were analyzed). Segments were rejected based on HAPPE’s recommended amplitude cut off (40μV) and joint-probability rejection criteria. 55 randomly selected segments were chosen for each participant for analysis.

### EEG rejection criteria

Participants were excluded if they had fewer than 55 EEG segments of usable data or fell outside 3 standard deviations from the mean on the following HAPPE quality control parameters: mean retained artifact probability (excluded versus included recordings: .239 versus .149), median retained artifact probability (.239 versus .113), percent of independent components rejected as artifact (63.5% versus 42.3%), and percent of EEG signal variance retained after artifact removal (28.2% versus 58.8%). Based on the above EEG rejection criteria, 13 of the 92 EEGs collected were rejected (11 RTT participants and 2 TD).

We note that included and excluded participants were similar in age (mean ± SD: 8.76 ± 6.39 and 9.69 ± 6.94 years, respectively) and in their distributions across studies and acquisition sites. Excluded RTT participants had similar clinical severity scores compared to included RTT participants (22.64 ± 4.50 versus 20.02 ± 7.31), and similar distributions of seizure frequency: no current seizures (54.5% versus 54.8%), seizures controlled with medication (18.2% versus 23.8%), monthly seizures (18.2% versus 11.9%), weekly seizures (9.1% versus 7.1%), and daily seizures (0% versus 2.4%).

Participant characteristics and all other data are reported for participants included in final analysis only.

### Phase-amplitude coupling

As previously described in Mariscal (70) and Peck (28), the following PAC computations were computed using the *pactools* Python toolbox. For each segment, first narrowband neural oscillations at low (2-20 Hz in 2 Hz steps) and high (20-100 Hz in 4 Hz steps) frequencies were extracted. Low frequencies were filtered using a constant bandwidth of 2 Hz, while high frequencies were filtered using an upper sideband variable bandwidth, equal to the low frequency of interest (71). Filtering at this step consisted of a zero-phase cosine-based filter to extract the real component and then a sine-based filter to extract the imaginary component, resulting in a complex-valued output signal (72).

The low frequency phase and high frequency amplitude time series were obtained from the filtered signals. The phases of the low frequency signals were binned into 18 20-degree intervals (-180° to 180°), and the mean of the amplitude of the high frequency signals within each phase bin was calculated. The amplitude values were then normalized by dividing each bin average by the sum of all bin averages. The resulting phase-amplitude distributions were averaged across all 55 segments within participant to minimize noise.

After averaging the phase-amplitude distribution across segments, coupling strength, also known as the Modulation Index (MI*raw*), was computed as the Kullback-Leibler divergence of the phase-amplitude distribution from a uniform distribution (73). To isolate the effect of PAC strength from related factors that may generate spurious phase-amplitude coupling (e.g. spectral power), phase amplitude distributions and coupling strength were also computed from a surrogate dataset that did not represent real effects of PAC. This surrogate dataset (MI*surr*) was generated by randomly circularly shifting low-frequency phase relative to high-frequency amplitude, with shifts ranging from 0.1 seconds to the signal duration minus 0.1 seconds. The modulation index was computed for 200 different iterations of this surrogate, null phase-amplitude coupling data. Next, a normalized z-scored modulation index was computed using the surrogate distribution; this method was proposed in Dupré La Tour (72). In carrying out these steps three measures were obtained from the original PAC strength: MI*raw*, MI*surr*, and MI*norm*, where MI*norm* is computed as the z-score of the MI*raw* compared to the distribution of MI*surr* values (previously termed z-MI in Mariscal (70)).

### Phase-amplitude coupling: all frequencies and regions

To characterize differences in patterns of PAC agnostic to predefined frequency bands, a data-driven approach was first employed, where analysis was performed across all frequency pairs and all channels.

To identify which frequency pairs and channels had statistically significant PAC within each group, a cluster-based approach was utilized, similar to Mariscal (70). The following steps were repeated for the RTT and TD groups separately.

1. For each low and high frequency pair within every channel, a t-test was conducted to compare the MI*raw* values across all participants within the group to the mean MI*surr* values across all participants within the group.
2. “Frequency grouping” size was identified as the number of adjacent significant frequency pairs within a channel, excluding diagonals.

To determine the significance of these frequency grouping sizes, MI*raw* and MI*surr* were flipped in a randomly selected subset of half the participants ranging from 1 to half the sample, and the same test statistics and frequency grouping sizes were computed (steps 1 and 2 above). This process was repeated 1000 times to generate a null distribution of frequency grouping size. Finally, the original frequency grouping sizes that were larger than the 95th percentile of this null frequency grouping size distribution were determined to reflect significant PAC.

To determine differences in PAC strength (MI*norm*) between groups, a cluster-based approach was utilized again, following the same general procedure described above. For each low- and high-frequency pair within every channel, an independent-samples t-test was conducted to compare MI*norm* between the RTT and TD groups (rather than MI*raw* and MI*surr* from all participants). Frequency grouping size was defined as the number of adjacent significant frequency pairs (*P* < 0.05) within a channel, excluding diagonal connections. To determine the significance of these frequency grouping sizes, participant group labels were randomly shuffled while preserving the original group sizes, and the same test statistics and frequency grouping sizes were recomputed. This process was repeated 1,000 times to generate a null distribution of frequency grouping sizes. Observed frequency groupings larger than the 95th percentile of this null distribution were considered to reflect statistically significant differences in PAC strength between groups.

### Primary Analysis: Phase-amplitude coupling strength in predefined regions of interest and frequency pairs

Next, predefined regions of interest and frequency pairs were examined. Group differences in PAC strength were evaluated using participant-level surrogate-normalized modulation indices (MI*norm*). PAC strength was calculated across four brain regions of interest: anterior (FP1, FP2, F3, F4, F7, F8, Fz), posterior (O1, O2, T5, T6, Pz), midline (C3, C4), and whole-brain (all 10-20 channels), and in the following frequency pairs: theta (4-8 Hz) and gamma (28-56 Hz) and alpha (9-13 Hz) and gamma (28-56 Hz). For each participant, MI*norm* values were averaged across the selected frequency bins and electrodes within each region. A series of eight independent-samples t-tests were used to test for differences between groups. Benjamini–Hochberg false-discovery-rate correction was applied across these eight primary PAC strength comparisons, with *q* < 0.05 considered significant. Effect sizes were reported as Hedges’ *g* with 95% confidence intervals.

### Secondary Analysis: Covariate Adjustments and descriptive analyses

Age was compared between the RTT and TD groups using an independent-samples t-test. Secondary linear regression analyses examined age and spectral power as potential covariates of each of the eight primary PAC strength outcomes. Age models included group, the main effect of age and the group-by-age interaction. Power spectral density (PSD) was estimated using multitaper analysis and averaged within each frequency band, across segments and corresponding electrodes used for PAC. Power models included group, log10-transformed PSD from the corresponding region and frequency bands (theta and gamma power for theta–gamma PAC; alpha and gamma power for alpha–gamma PAC), and group-by-power interactions. Age and power were evaluated in separate models to avoid overfitting.

To determine whether the primary group differences remained after covariate adjustment, group effects were estimated in models adjusted separately for age and power. Benjamini–Hochberg FDR correction was applied across the 16 adjusted group comparisons: eight from the age-adjusted models and eight from the power-adjusted models.

Within the RTT group, clinical severity was assessed by clinicians using the Clinical Severity Scale (CSS) (74) and Clinical Global Impressions of Severity (CGI-S) (75, 76). Correlations were used to examine the association between CSS or CGI score and each of the eight primary PAC strength outcomes. Participants were classified as having no seizures and not receiving antiseizure medication (n = 23), no seizures while receiving antiseizure medication (n = 10), or active seizures (n = 9; monthly, n = 5; weekly, n = 3; daily, n = 1). PAC strength was summarized across these seizure frequency categories. Medical history and medication use were recorded, and medications were reviewed in relation to PAC strength. To assess whether the findings were driven by a particular study or recording site, PAC-strength differences were also examined within study and site subsets. Given limited subgroup sizes, these analyses were not adequately powered and were therefore interpreted descriptively.

Clinical severity scores, seizure history, and other participant characteristics are reported in Table 1.

### Modeling PAC

We constructed a biophysical model of Layer 4 cortical networks using single-compartment neurons with Hodgkin-Huxley-type dynamics (Fig. 3A). The network model included excitatory pyramidal neurons or stellate neurons (PYR) and inhibitory interneurons. The interneuron populations consisted of the three most common types of interneurons found in cortex: PV+INs, SOM+INs and 5HT3aR+INs (77). We used the VIP+IN subtype of 5HT3aR+ INs in our model since we were particularly interested in how VIP+IN activity affects network dynamics—VIP+INs have been implicated in mediating Rett symptoms by specific deletion of MeCP2 in VIP+ cells (40). The model VIP+INs had D-, M- and T-currents which allowed for bursts of spikes at a delta time scale. L4-targeting SOM+INs (also known as X94 SOM+ interneurons) have a unique electrophysiological profile among cortical SOM+ cells, which includes stuttering behavior at alpha frequency (40, 78). Our model L4 SOM+INs had D-, A- and h-type membrane currents that produced the alpha frequency stuttering. We note that rodent L4 differs in cell types and circuits depending on cortical area. We chose rodent S1—the cortical areas mediating the rodent’s specialized sense of somatosensation—to model human L4, which includes X94 SOM+ cells. The model was intended to test a candidate circuit mechanism rather than to reproduce all anatomical and biophysical properties of human cortex. The PV+INs and PYRs were modeled using previous formulations of these cell types (79). Each cell type was connected to other types of neurons and to other cells of its own type with varying types, strengths and degrees of connectivity constrained by existing literature. The network consisted of 110 neurons, with the number of each cell type roughly proportional to their prevalence in cortex (77). All neuronal subtypes were connected to each other through excitatory (AMPA) or inhibitory (GABAa) synapses, with weights assigned according to available information. All neurons within the same class were connected to each by electrical and/or chemical synapses.

We studied the network activity under typical and low VIP+IN activity levels, running 20 simulations for each condition. To decrease the activity of VIP+INs we lowered the maximal sodium current conductance of the VIP+INs to 80% of its baseline value, which lowered the spiking rate of VIP+INs but did not silence them. To examine the effect of lowered levels of activity of other neuronal subtypes in the model, we lowered each subtype’s maximal sodium current conductance to 80% of its baseline level.

The local field potential (LFP)—similar to EEG in that it records extracellular voltage fluctuations but different in that it is more spatially restricted—was modeled as the sum of all excitatory currents among PYR neurons in the network (80). To compute PAC we followed the methods in Kramer et al., 2016 (81). PAC was determined by first bandpass filtering the model LFP at gamma, theta and alpha frequencies; the range of theta was 4 – 8 Hz, the range of alpha was 9 – 13 Hz and the range of gamma was 20 – 60 Hz. This was followed by extraction of phase and amplitude information of low (theta and alpha) and high (gamma) frequencies using the Hilbert transform; this procedure generated phase-amplitude time-series vectors. We then computed the average gamma amplitude for each phase of the low frequency oscillations and found the difference (h) between the maximum and minimum values of the average amplitude. To determine statistical significance of h, we compared h to the difference between the maximum and minimum values of h produced from 1000 surrogate phase-amplitude vectors generated by randomly permuting the amplitude time series to assign each time point of the phase vector to a random amplitude. Significant PAC was determined by computing the proportion of h-values from the surrogate phase-amplitude data greater than the h-value from the non-randomized phase-amplitude data (81). The normalized modulation index (MI) for each high and low frequency pair was calculated using a similar method to the one described above (73), and a two-sample t-test was used to evaluate significant differences between MI for simulations run under different conditions.

The code for the model was written in C++; data analysis and graphics used MATLAB. See SI Appendix, Computational Methods for more details.

## Results

To investigate brain-wide oscillatory dynamics in RTT, we analyzed phase-amplitude coupling during resting awake-state EEG recorded from 42 individuals with a confirmed diagnosis of RTT and 37 typically developing participants. We obtained low frequency phase and high frequency amplitude time series from the EEG data and extracted phase-amplitude distributions, used throughout the following analyses (see Methods).

### PAC in Rett syndrome and typical development across all possible frequencies and regions

We used a data-driven approach agnostic to predefined frequency ranges to explore any patterns in PAC that may not be captured otherwise, evaluating coupling between all possible frequency pairs and across all 18 electrodes.

Because PAC has not been previously described in the awake state in RTT, we first asked at what channels and at what frequency pairs significant PAC occurs. Using a cluster-based permutation analysis, we compared observed PAC to randomized data to identify regions and frequency pairs where coupling was significant. Fig. 1A displays instances of significant coupling for RTT and TD independently, in a binary manner, agnostic to the strength of the coupling. Significant PAC was observed at all 18 electrodes in both groups, with substantial overlap across frequency pairs (Fig. 1A).

**Figure 1.**
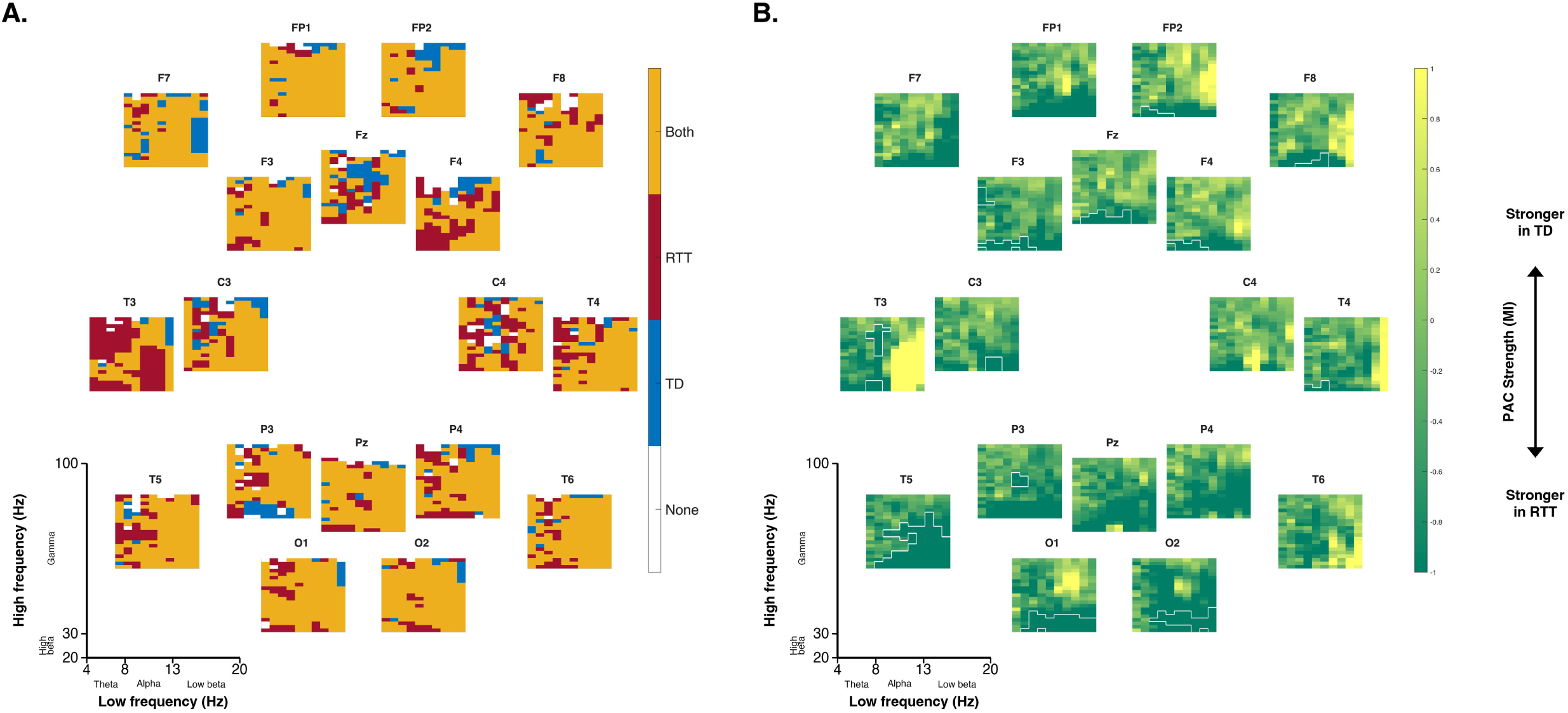
Phase-amplitude coupling in RTT and TD across all possible frequencies and regions. Comodulograms arranged by scalp position for all 18 electrodes of the 10-20 channel array for RTT (n = 42) and TD (n = 37). Each subplot represents one electrode, and all subplots share the same axes of low and high frequency filtered signal, as shown in the lower left corners. **(A)** Instances of significant PAC frequency pairs shown for RTT and TD independently. Significant pairs are color-coded for RTT (red) and TD groups (blue), with any overlapping significant coupling shown in orange. Within every subplot, each (*x*, *y*) coordinate with colored shading represents significant coupling between the corresponding low frequency (*x)* and high frequency (*y)* signals. Significance was determined using a cluster-based permutation procedure. **(B)** Difference in mean normalized PAC strength (MI*norm*) between RTT and TD. On every subplot, each (*x*, *y*) coordinate represents RTT subtracted from TD strength of coupling between the corresponding low frequency *x* and high frequency *y* signal. Dark green shading corresponds to higher PAC strength in RTT than TD, whereas bright yellow shading corresponds to higher PAC strength in TD than RTT. Lighter green indicates areas of low PAC strength difference. White boundaries outline areas where coupling strength is significantly different between groups; significance was calculated using a cluster-based permutation procedure.

To investigate differences in PAC between RTT and TD using this data-driven approach, we next quantified the *strength* of coupling, comparing normalized modulation index (MI) values between groups. In Fig. 1B, RTT mean MI was subtracted from TD and plotted to visualize the difference in coupling strength between groups. The direction of these differences predominantly indicated stronger PAC in RTT compared to TD in a large portion of frequency pairs and electrodes. Using a cluster-based permutation analysis, we then compared observed group differences with randomized group assignments and identified frequency pairs where PAC strength was significantly different between groups in 12 of the 18 electrodes, all in the direction of higher PAC strength in RTT (Fig. 1B). These differences predominantly involved slower frequency phase coupled to lower-gamma amplitude.

### PAC strength is increased in Rett syndrome in theta-gamma and alpha-gamma frequency pairs in primary analysis

We next quantified coupling strength for each participant within the groups, focusing on four regions of interest (anterior, posterior, midline, and whole-brain) and two predefined frequency pairs that have been most affected in other neurodevelopmental disorders (alpha-gamma and theta-gamma) (28, 33, 70, 71). We found that RTT participants had significantly higher alpha-gamma MI compared to TD in anterior, posterior, and whole-brain regions, and significantly higher theta-gamma MI in posterior, midline, and whole-brain regions (independent-samples t-tests; all *q* < 0.05; Fig. 2, Table 2). Effect sizes ranged from moderate to large (Hedges’ *g* = 0.53–0.91; Table 2). While individual values overlapped between groups (a subset of participants in the RTT group showed MI values comparable to those in the TD group), the distributions were shifted toward higher PAC strength in RTT in all regions we evaluated (Fig. 2).

**Figure 2.**
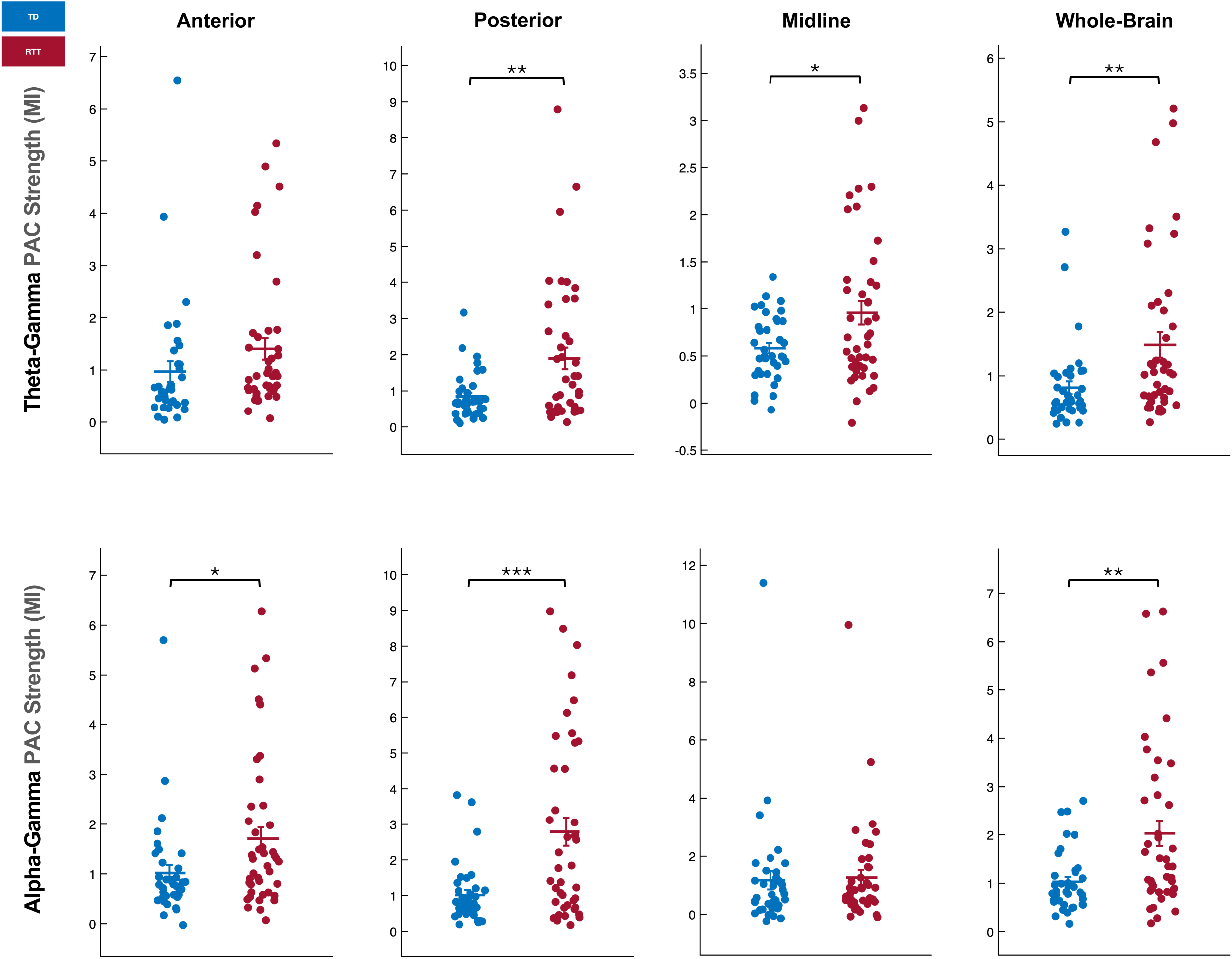
Theta-gamma and alpha-gamma PAC strength elevated in individuals with Rett syndrome. Markers in each subplot signify a single individual’s normalized PAC strength for a given frequency pair and region (indicated on the top and left). TD participants (*n* = 37) are shaded in blue and RTT participants (*n* = 42) are shaded in red. Horizontal bars and error bars show the group mean ± SEM. Group differences were evaluated using independent-samples t-tests with Benjamini–Hochberg FDR correction across eight comparisons, where \**q* < 0.05, \*\**q* < 0.01, and \*\*\**q* < 0.001.

**Table 2.** PAC strength comparison in predefined frequency pairs.

|  |  | **Diagnostic Group** |  |  |  |  |
|---|---|---|---|---|---|---|
| **Outcome** | **Region** | **RTT (n = 42)** | **TD (n = 37)** | **Effect size [95% CI]** | **p** | **FDR q** |
| **PAC strength, theta-gamma** | Whole brain | 1.49 ± 1.29 | 0.81 ± 0.62 | g = 0.65 [0.19, 1.10] | **0.005** | **0.01** |
|  | Anterior | 1.40 ± 1.34 | 0.97 ± 1.20 | g = 0.34 [-0.11, 0.78] | 0.136 | 0.156 |
|  | Posterior | 1.90 ± 1.94 | 0.85 ± 0.63 | g = 0.70 [0.25, 1.16] | **0.002** | **0.006** |
|  | Midline | 0.96 ± 0.80 | 0.58 ± 0.34 | g = 0.59 [0.14, 1.04] | **0.01** | **0.016** |
| **PAC strength, alpha-gamma** | Whole brain | 2.03 ± 1.71 | 1.03 ± 0.63 | g = 0.75 [0.29, 1.21] | **0.001** | **0.005** |
|  | Anterior | 1.71 ± 1.50 | 1.02 ± 0.97 | g = 0.53 [0.08, 0.98] | **0.02** | **0.026** |
|  | Posterior | 2.79 ± 2.54 | 1.02 ± 0.84 | g = 0.91 [0.44, 1.37] | **<.001** | **<.001** |
|  | Midline | 1.26 ± 1.73 | 1.18 ± 1.95 | g = 0.05 [-0.40, 0.49] | 0.837 | 0.837 |

Values are mean +/- SD. PAC strength was tested with equal-variance independent-samples t tests. g = Hedges g. Benjamini-Hochberg FDR correction was applied across the 8 PAC-strength comparisons; bold q values are < .05.

Unless otherwise specified, subsequent references to PAC strength refer to these primary PAC strength outcomes rather than the exploratory data-driven analysis.

### PAC strength differences remain consistent across secondary analyses

#### Age and spectral power

Age did not differ significantly between the RTT and TD groups (*t*(77) = 0.60, *P* = 0.550; Table 1), but because PAC can vary across development, we examined age as a potential covariate of PAC strength (82). In addition, although PAC is a distinct and standalone measure, given that spectral power is a more commonly assessed feature of EEG, secondary models adjusted PAC strength differences for power in the corresponding frequency bands and regions. Neither age nor spectral power, nor their interaction with group, significantly predicted PAC strength. We then tested whether the primary group differences remained in models adjusted separately for age and spectral power. All primary PAC strength differences remained significant after adjustment for age (Table S1). Participant-level relationships between age and PAC strength are shown in Fig. S1. Five of six primary differences remained significant after adjustment for spectral power; anterior alpha–gamma PAC remained higher in RTT but did not meet significance (*q* = 0.071; Table S1). Participant-level relationships between power and PAC strength are shown in Figs. S2 and S3.

#### Clinical characteristics

Within the RTT group, correlations did not indicate a consistent relationship between clinical severity and PAC strength. PAC strength also showed no systematic variation across seizure-frequency categories, and review of medication use revealed no discernible pattern between medication use and PAC strength (Table S2).

#### Study and recording site

PAC strength differences were generally consistent in direction across study and site subsets compared to those observed in the combined sample, although statistical support was weaker in the smaller subsets.

### VIP+IN dysfunction increased theta-gamma and alpha-gamma PAC strength in a Layer 4 network model

To explore the underlying cellular and network mechanisms of changes to PAC in RTT, we constructed a biophysically constrained model of cortical Layer 4 containing excitatory neurons (PYR) as well as PV+, SOM+ and VIP+ interneurons (Fig. 3A). Since VIP+INs have been implicated in mediating RTT symptoms (40), we were particularly interested in whether deficiency in VIP+IN activity could account for the elevated PAC we observed in RTT. We therefore studied the model network activity under typical (baseline) and low VIP+IN activity levels.

**Figure 3.**
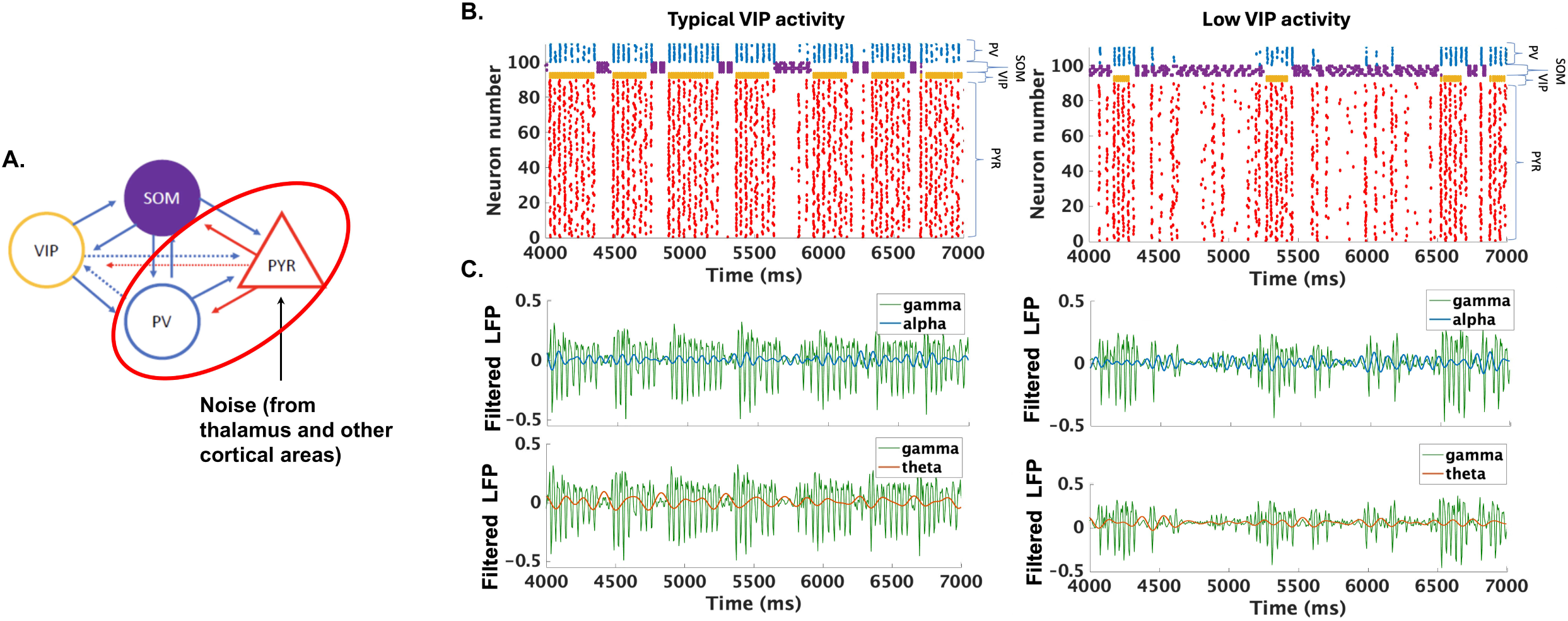
VIP+ interneuron hypoactivity alters local network dynamics but PAC remains significant in L4 model. **(A)** Schematic of L4 network. PYR represents excitatory stellate and/or pyramidal cells, PV represents fast spiking parvalbumin+ cells, SOM represents somatostatin+ cells projecting to L4, and VIP represents VIP+ cells. Each symbol represents a population of cells of a given cell type. In isolation (without network connections), VIP+ cells produce delta/theta (2 – 4 Hz), SOM+ cells produce alpha, and the reciprocal interaction between PYR and PV+ cells produces gamma via PING (pyramidal cell – interneuron gamma). Noisy input is given to pyramidal cells to represent thalamic input and/or input from other cortical areas. Excitatory synaptic connections are represented by red arrows and inhibitory connections are represented by blue arrows. **(B)** Raster plots of simulations of L4 network under conditions of typical VIP+IN activity and when VIP+IN activity is lowered. **(C)** Filtered LFP signals show high (gamma in green) and low (alpha in blue and theta in orange) frequency bands and their coordination under typical and low VIP+IN activity conditions. Significant PAC was assessed separately within each condition by comparing observed coupling with randomized surrogate data using a permutation test.

At typical (baseline) levels of VIP+IN activity, networks showed significant participation of all neuronal subtypes (Fig. 3B, left). VIP+INs and SOM+INs were mutually inhibitory, and the network toggled between periods of SOM+IN-dominated activity and periods of VIP+IN-dominated activity. The brief periods of SOM+IN activity were accompanied by pauses in network activity, whereas the relatively longer VIP+IN activation periods allowed PYR and PV+INs spiking. Gamma oscillations emerged in the network spiking during periods of VIP+IN activity (Fig. 3B left, 3C left) when PYR and PV+INs interacted to form a type of gamma rhythm known as PING (pyramidal-interneuron network gamma) (83) as observed in the filtered model LFP (Fig. 3C, left). Although theta and alpha oscillations were also apparent, they were not clearly attributable to the spiking activity of any one neuronal subtype. Nevertheless, significant LFP PAC relative to randomized surrogate data was found in the baseline condition between both theta and gamma (*P* < 0.01) as well as between alpha and gamma (*P* < 0.05) (Fig. 3C, left).

We next considered simulations in which VIP+IN excitability was decreased. The overall spiking rate of PYR was greatly reduced (Fig. 3B, right), consistent with findings in mouse models of RTT (35). The reduction of activity was a consequence of overactivity of SOM+INs, which were released from baseline levels of VIP+IN inhibition, thereby strongly inhibiting all other network components. Although the population spiking of PYRs was decreased compared to baseline, gamma still emerged from PING oscillations, which occurred robustly during the brief VIP+IN active periods and sparsely during the periods of SOM+IN overactivity. Despite obvious changes in network dynamics (compared to baseline) when VIP+IN activity was low, the PAC between low frequency (theta and alpha) and high frequency (gamma) oscillations in the model LFP remained significant (*P* < 0.05 in both cases) (Fig. 3C, right). These results are consistent with our finding of significant theta-gamma and alpha-gamma coupling in the EEG of *both* TD and RTT participants as shown in Fig. 1A.

Importantly, if reduction in VIP+IN activity is related to the changes in PAC strength found in our EEG data, simulations in which VIP+IN activity is lowered should increase PAC coupling strength compared to the PAC strength under baseline conditions. Therefore, we next investigated if the *strength* of the gamma modulation by theta and alpha (coupling strength, or MI) changed when VIP+IN activity was lowered. When we compared baseline and low VIP+IN activity conditions, MI was significantly increased in the low VIP+IN condition in both theta-gamma (*P* < 0.01) and alpha-gamma (*P* < 0.05) (Fig. 4), matching the observations in the EEG of RTT participants as shown in Fig. 2.

**Figure 4.**
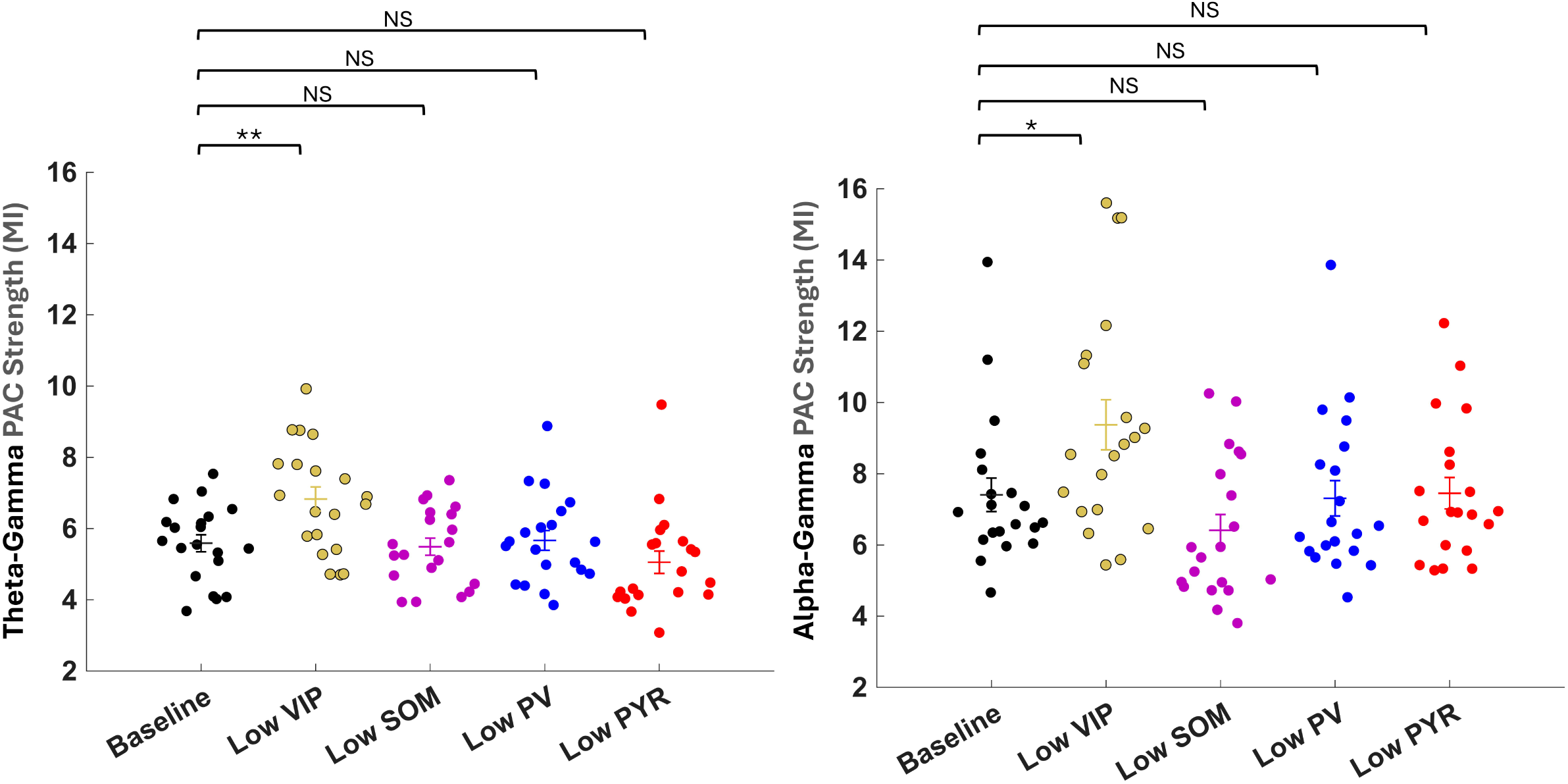
Increased modulation of gamma by slower theta and alpha frequencies with reduced VIP+ interneuron excitability. Markers represent the normalized modulation index (MI*norm*) of theta–gamma (left) or alpha–gamma (right) PAC in the model LFP after 9s of simulation time. Black markers represent typical (baseline) activity levels of all neurons in the network. Gold, purple, blue, and red markers represent low VIP+IN, SOM+IN, PV+IN, and PYR activity, respectively. n = 20 for each group. Horizontal bars and error bars show the group mean ± SEM. Each low-activity condition was compared with baseline using independent-samples t-tests, where \**P* < 0.05, \*\**P* < 0.01, and NS indicates not significant.

Finally, we probed whether other neuronal subtypes (PV, SOM or PYR) could account for some of the changes to PAC coupling strength seen in the RTT group by repeating the simulations and lowering the activity of one neuron subtype at a time. Lowering the activity of PV, SOM or PYR did not cause any significant changes in PAC compared to baseline (Fig. 4). Thus, VIP+IN activity emerged as a key component of changes to theta/gamma and alpha/gamma PAC in our model networks. Overall, our model identifies VIP+IN dysfunction as a candidate circuit mechanism that could contribute to the elevated PAC strength observed in RTT.

## Discussion

Rett syndrome is a disorder of widespread brain dysfunction, yet its impact on large-scale network activity remains incompletely understood. Oscillatory dynamics—specifically phase–amplitude coupling—offer a powerful, non-invasive window into these disruptions in RTT. With few disease-altering therapeutics currently available, such measures may be valuable for characterizing circuit dysfunction and supporting the development and evaluation of targeted interventions.

Here we investigated phase-amplitude coupling in the awake resting state in individuals with RTT compared to TD. We found that both groups exhibited significant PAC across all electrodes and a broad range of frequency combinations, confirming widespread cross-frequency coordination at rest, independent of any presence of disorder. PAC strength was broadly elevated in RTT, and analyses of predefined frequency bands further revealed increased theta–gamma PAC strength in posterior, midline, and whole-brain regions, and increased alpha-gamma PAC strength in anterior, posterior, and whole-brain regions. Finally, simulations from a biophysically constrained model showed that reduced excitability in VIP+ interneurons (but not the other neuronal subtypes tested) produced similar elevations in theta-gamma and alpha-gamma PAC strength, supporting a candidate mechanistic link between VIP+IN dysfunction and the empirical data. Together, these findings suggest that PAC captures systems-level abnormalities in neural coordination that may be driven by a particular cell type in RTT.

Elevated theta–gamma coupling in Rett syndrome is notable given the established role of theta-gamma coupling in working memory, episodic encoding, and cognitive control (23–26). In both humans and animal models, theta phase organizes gamma bursts into discrete representational windows, supporting flexible information integration (23–26). In the context of RTT, increased theta–gamma coupling may reflect compensatory recruitment of neural resources or inefficient top-down modulation. Alternatively, it could indicate disrupted thalamocortical coordination, a feature implicated in both RTT and other neurodevelopmental conditions (19, 41, 56, 84). Altered alpha–gamma coupling in RTT further implicates disruptions in sensory integration and attentional gating. Alpha rhythms modulate cortical excitability and control sensory gain, while gamma activity reflects local circuit computation (20, 85–87); increased alpha–gamma coupling may therefore reflect a compensatory mechanism aimed at stabilizing sensory processing through enhanced top-down control. Evaluating PAC metrics in event or task-related paradigms will further clarify the functional role of coupling during active cognitive or sensory engagement.

Among the neuronal perturbations tested in our computational model of L4, dysfunction in VIP+INs alone recapitulated the elevated theta–gamma and alpha–gamma coupling observed in the EEG data, suggesting that lowered VIP+IN activity could contribute to PAC abnormalities in RTT. Our computational results also suggest that VIP+IN deficiency alters network function by increasing SOM+IN modulated inhibitory tone, which restricts signal transmission that would typically occur with higher VIP+IN activity levels. To establish causality between deficiency of VIP and increased theta/gamma and alpha/gamma coupling in RTT, future studies could directly manipulate VIP+IN function in RTT preclinical models. Selective deletion of *Mecp2* from VIP+INs followed by *in vivo* EEG recording would test whether *Mecp2* loss-induced VIP+IN dysfunction is sufficient to produce elevated PAC. If causality is established between deficiency of VIP+INs and increased theta/gamma and alpha/gamma coupling, future studies can combine experimental event or task-related paradigms with further modeling to identify functional consequences of the altered PAC. Complementary experiments restoring VIP+IN function—either genetically or through targeted neuromodulation—could provide critical insight into reversibility. Notably, non-invasive stimulation approaches that entrain gamma rhythms have been shown to improve pathology via VIP peptide (a peptide co-released with GABA from VIP+INs) in a mouse model of Alzheimer’s disease, and may offer a translatable strategy to modulate VIP+IN activity in RTT (88).

We observed inter-individual variability in PAC strength within the RTT group, with some participants exhibiting MI values similar to TD and a subset exhibiting higher values. Chronological age did not significantly predict PAC strength, with all primary group differences remaining significant after age adjustment. Disease stage may nevertheless contribute to interindividual variability in PAC strength, potentially reflecting distinct states of underlying circuit pathology. Additionally, the type or location of *MECP2* mutations, as well as differential patterns of X chromosome inactivation skewing (on a cell-by-cell and regional basis), influence disease severity (74, 89–92) and could thus impact the nature of PAC alterations and impairment to VIP+INs. Interestingly, similar variability in PAC strength was observed in the model under low VIP+IN conditions (where the only source of variability between individual model simulations was noise), suggesting that low VIP+IN activity predisposes the network to increased PAC strength under certain noise conditions. Noise in our model corresponds to undefined background processing states of the brain, which could parallel different cognitive or behavioral states in individuals. Thus, our model suggests that inter-individual variability could also be brain-activity dependent and determined by the cognitive processes active at the time the EEG is recorded. Preclinical and clinical studies in RTT systematically studying distinct stages of disorder, genetic subtypes, and behavioral states will help clarify inter-individual variability in electrophysiological phenotypes.

Several additional limitations should be considered. Although comparatively large for a rare genetic disorder, the sample remains modest even after combining data across studies. Despite robust differences between the RTT and TD groups, PAC strength showed no consistent relationship with clinical severity within the RTT group; therefore, its relationship to individual differences in clinical presentation remains uncertain. Study/site subsets and seizure and medication subgroups were small, limiting definitive evaluation of heterogeneity related to these factors. Because PAC was measured during a single awake recording, its within-person stability across recording sessions and behavioral states remains unknown, and the cross-sectional design prevents us from determining how PAC changes across development. Longitudinal studies and independent samples could help determine whether PAC alterations arise early or change with ongoing network pathology and establish their reliability, specificity, and sensitivity to disease progression or treatment. The primary group differences in PAC strength were largely preserved after accounting for spectral power; however, power adjustment, surrogate normalization, and standardized artifact removal reduce but do not eliminate the possibility of other influences or confounds on PAC. Finally, our Layer 4 model and model-derived LFP do not directly reproduce human scalp EEG; rather, they identify reduced VIP+ interneuron excitability as one plausible mechanism for increased PAC at the local circuit level.

Overall, our findings support the potential of PAC as an objective, non-invasive marker of inhibitory circuit dysfunction. Reports of disrupted theta–gamma coupling in Fragile X syndrome and ASD (31, 33) and altered alpha–gamma phase bias in Phelan–McDermid syndrome (70) and ASD suggest that aberrant PAC may reflect shared network-level vulnerabilities across neurodevelopmental disorders, despite divergent genetic etiologies. PAC may therefore provide a systems-level framework for comparing circuit dysfunction and identifying mechanistically meaningful subgroups across neurodevelopmental and neuropsychiatric disorders (31, 33). Such work could advance clinical-trial readiness by supporting biologically informed participant stratification and the development and testing of interventions targeting shared inhibitory circuit dysfunction. As our understanding of cell-type-specific contributions to oscillatory dynamics grows, PAC may help connect genetic and cellular pathology with systems-level neurophysiological outcomes and, ultimately, therapeutic response across a broad range of neurological disorders.

## Data Availability

Data are available upon reasonable request from both NHS and RSBS studies. Code used for modeling will be made available on GitHub.

## Author Contributions

DK, AL, and MF conceptualized the study. DK wrote and prepared the manuscript draft; YB contributed to the PAC Methods section and MM to the model sections. DK performed the analysis. YB helped develop analysis scripts, and MM provided the main computational contribution. DK and KS collected clinical data for the Rett Syndrome Biomarkers Study. EDM, AP, JN, and JS oversaw and collected clinical data for the Rett and Related Disorders Natural History Study. AL, MF, and CN supervised the project. All authors contributed to interpretation of the results and critical revision of the manuscript.

## Acknowledgements

Thank you very much to all the participants in these studies and their families, especially those who travelled and extended themselves to contribute to research in Rett syndrome. Thank you to the Rett Syndrome Angels (originally Rett Syndrome Association of Massachusetts) for their incredible support. Thank you to the Rett syndrome clinics and to all members of the labs of Dr.’s April Levin, Michela Fagiolini, Charles Nelson, and Takao Hensch for your invaluable feedback and contributions. Thank you to all the sites and people that participated in the Rett and Related Disorders Natural History Study. This work could not have been completed without all of you.

## Funding

This work was supported partially by:

P50HD105351 – Boston Children’s Hospital Intellectual and Developmental Disabilities Research Center – Project 1 – Michela Fagiolini

3U54HD061222 – For Rett syndrome, MECP2 Duplications, and Rett-related Disorders

Natural History – Alan Percy, Eric Marsh, Tim A. Benke, Jeffrey Neul

P50HD103537 – Jeffrey Neul

The University of Tokyo International Research Center for Neurointelligence (WPI-IRCN) – Takao Hensch

