## Supplementary material for "Altered oscillatory coupling and a candidate inhibitory circuit mechanism in Rett syndrome": SI Appendix

#### Index

Computational Methods S2

SI References S12

Tables S1–S2 S14

Figures S1–S3 S17

#### Computational Methods

##### *Neurons*

Our model of the cortical Layer IV uses single-compartment, biophysical models of four types of neurons: PV+INs, VIP+INs, SOM+INs and excitatory cells (pyramidal cells or stellate cells) called PYR. The voltage ($V$) of each neuron changes in time according to an equation that equates the capacitive membrane current of each neuron with its membrane ionic currents. This equation is given by:

$$c_m \frac{dV}{dt} = -\sum I_{memb} - \sum I_{syn} + I_{app} \tag{1}$$

In (1), the membrane voltage ($V$) has units of $mV$. The membrane ionic currents ($I_{memb}$), the synaptic current ($I_{syn}$), and the applied current ($I_{app}$) all have units of $\mu A/cm^2$. The specific membrane capacitance ($c_m$) is set to $1 \mu F/cm^2$ for all neurons. All ionic membrane and synaptic current have Hodgkin-Huxley-type conductances formulated as:

$$I = \bar{g} m^n h^k (V - E_{ion}) \tag{2}$$

In (2), the maximal ionic conductance ($\bar{g}$) uses units of $\mu S/cm^2$ and the ionic reversal potential ($E_{ion}$) has units of $mV$. Each ionic current has $n$ activation gates ($m$) and $k$ inactivation gates ($h$). For the membrane currents, the activation and inactivation gating variables evolve according to a two-state kinetic equation formulated (written for the gating variable $m$) as:

$$\frac{dm}{dt} = \frac{m_\infty(V) - m}{\tau_m(V)} \tag{3}$$

The steady-state function ($m_\infty(V)$) and the time constant of decay ($\tau_m(V)$) are dependent on voltage. These functions may be re-written as rate functions ($\alpha_m(V), \beta_m(V)$) using the transformation:

$$m_\infty(V) = \alpha_m(V)/(\alpha_m(V) + \beta_m(V))$$
$$\tau_m(V) = 1/(\alpha_m(V) + \beta_m(V))$$

The equations for the synaptic currents ($I_{syn}$) describe the dynamics of the interaction between the presynaptic (*pre*) and postsynaptic (*post*) neurons:

$$I_{post} = \bar{g}s(V_{pre})(V - E_{ion}) \tag{4}$$

Each post synaptic current has a single activation gate ($s$), which is dependent on the voltage of the pre-synaptic neuron ($V_{pre}$). The applied current ($I_{app}$) is either a constant term or a noise term or a sum of both. $I_{app}$ determines the excitability level of an individual cell.

***Excitatory Cells***

The excitatory neurons have spiking currents (i.e., sodium current ($I_{Na}$), potassium current ($I_K$) and leak current ($I_L$)) as well as an M-current ($I_M$) and an h-current ($I_h$), both known to be found in cortical pyramidal cells (1, 2). Synaptic currents to the excitatory cells come from inhibitory input from SOM+INs ($I_{som_e}$), PV+INs ($I_{pv_e}$) and VIP+INs ($I_{vip_e}$) as well as excitatory input from other excitatory cells ($I_{e_e}$). The voltage change in an excitatory cell is described by:

$$c_m \frac{dV}{dt} = -I_{Na} - I_K - I_L - I_M - I_h - I_{pv_e} - I_{vip_e} - I_{som_e} - I_{e_e} + I_{app} \tag{5}$$

Models of the spiking currents ($I_{Na}, I_K, I_L$) are taken from a previous formulation of these currents (3). The maximal sodium conductance is set to $\bar{g}_{Na} = 100\, mS/cm^2$. The sodium reversal potential is $E_{Na} = 50mV$. This sodium current has three activation gates ($n = 3$) and one inactivation gate ($k = 1$). The rate constants for the activation ($m$) and inactivation ($h$) variables are described by:

$$\alpha_m = \frac{0.32(V+54)}{1-\exp[-(V+54)/4]}$$
$$\beta_m = \frac{0.28(V+27)}{\exp[(V+27)/5]-1}$$
$$\alpha_h = 0.128\exp[-(V+50)/18]$$
$$\beta_h = \frac{4}{1+\exp[-(V+27)/5]}$$

The maximal conductance for the fast potassium channel is $\bar{g}_K = 80\, mS/cm^2$ and the reversal potential for potassium is $E_K = -100mV$. The fast potassium channel has no inactivation gates and four activation gates described by the rate functions:

$$\alpha_m = \frac{0.032(V+52)}{1-\exp[-(V+52)/5]}$$
$$\beta_m = 0.5\exp[-(V+57)/40]$$

The leak current ($I_L$) has no gating variable. The maximal leak channel conductance is $g_L = 0.1\, mS/cm^2$. The leak current reversal potential is $E_L = -67mV$.

The formulation of the M-current comes from Mainen et al., 1996 (4). It has one activation gate and no inactivation gate. The maximal conductance for the M-current, $\bar{g}_M$ is 1.3 mS/cm$^2$. The rate functions for the M-current activation variable are:

$$\alpha_m = \frac{Q_s 10^{-4}(V+30)}{1-\exp\left[-(V+30)/9\right]}$$
$$\beta_m = -\frac{Q_s 10^{-4}(V+30)}{1-\exp\left[(V+30)/9\right]}$$

For the M-current, a $Q_{10}$ factor of 2.3 is used in scaling the rate functions because the kinetics were originally derived from experiments performed at 23 °C. To model dynamics of the M-current at a normal body temperature of 37 °C, the rate equations for the M-current are scaled by:

$$Q_s = Q_{10}^{(37\,^\circ\mathrm{C}-23\,^\circ\mathrm{C})/10} = 3.209$$

The model for the h-current comes from Jones et al., 2000 (5). It has one activation gate and no inactivation gate. The maximal conductance for the h-current ($\bar{g}_h$) is 1.2 mS/cm$^2$. The reversal potential for the h-current is $E_h = -43mV$. The h-current activation variable ($r$) steady state function and time constant of decay are given by:

$$r_\infty(V) = \frac{1}{1+\exp((V+75)/5.5}$$
$$\tau_r = \frac{1}{\exp(-14.59-0.086\cdot V)+\exp(-1.87+0.0701\cdot V)}$$

The background excitation to each pyramidal cell was provided by giving independent Gaussian noise to each neuron with an amplitude of $120\cdot\sqrt{0.01}$ where 0.01 is the time step of integration. Network properties including synaptic currents ($I_{pv_e}, I_{vip_e}, I_{som_e}, I_{e_e}$) are discussed below.

***PV Cells***

Each PV neuron derives its membrane currents from the interneuron model in Olufsen et al., 2003 (3):

$$c_m\frac{dV}{dt} = -I_{Na} - I_K - I_L - I_{vip_{pv}} - I_{som_{pv}} - I_{pv_{pv}} - I_{e_{pv}} - I_{elec} \quad (6)$$

The formulations of the $I_{Na}$, $I_K$ and $I_L$ are the same as for our model excitatory cell including the same values for all maximal conductances and reversal potentials. The PV cells did not have a background excitation term and thus were silent without excitatory synaptic input. Electrical synapses ($I_{elec}$) and ionic synaptic connections ($I_{vip_{pv}}$, $I_{som_{pv}}$, $I_{pv_{pv}}$, $I_{e_{pv}}$) are discussed below.

***SOM Cells***

Our model SOM cells were constructed to mimic the dynamics of Layer IV projecting SOM cells (also known as X94 cells) as described in Ma et al., 2006, which include delayed spiking, stuttering (which can be at alpha frequency) and tonic spiking with increasing levels of depolarizing currents, as well as rebound excitation in response to hyperpolarizing currents (6). These dynamics were captured by inclusion of the spiking currents (sodium, potassium and leak), an A-type potassium current (for delayed spiking), a D-type potassium current for stuttering dynamics (7), and an h-current for rebound to hyperpolaization as suggested by Ma et al., 2006. Our SOM cells are formulated as:

$$c_m \frac{dV}{dt} = -I_{Na} - I_K - I_L - I_A - I_D - I_h - I_{vip_{som}} - I_{pv_{som}} - I_{e_{som}} - I_{elec} + I_{app} \quad (7)$$

The formulations of the $I_{Na}$, $I_K$ and $I_L$ derive from Golomb et al., 2007 (7). The maximal sodium conductance is set to $\bar{g}_{Na} = 112.5\,mS/cm^2$. The sodium reversal potential is $E_{Na} = 50mV$. The sodium current has three activation gates and one inactivation gate . The activation variable ($m$) is modeled by its the steady state function ($m = m_\infty$) formulated as:

$$m_\infty(V) = \frac{1}{1+\exp(-(V+24)/11.5)}$$

The kinetics of the inactivation gate ($h$) are given by:

$$h_\infty(V) = \frac{1}{1+\exp((V+58.3)/6.7)}$$

$$\tau_h = 0.5 + \frac{14}{1+\exp((V+60)/12)}$$

The fast potassium current is modeled using two activation gates and no inactivation gates. The maximal conductance of the fast potassium channel is $\bar{g}_K = 225\,mS/cm^2$. The potassium reversal potential is $E_K = -90mV$. The kinetics of the activation gate ($n$) are described by:

$$n_\infty(V) = \frac{1}{1+\exp(-(V+12.4)/6.8)}$$

$$\tau_n = \left[0.087 + \frac{11.4}{1+\exp((V+14.6)/8.6)}\right] \cdot \left[0.087 + \frac{11.4}{1+\exp(-(V-1.3)/18.7)}\right]$$

The leak current has maximal conductance $\bar{g}_L = 0.27\, mS/cm^2$ and its reversal potential is $E_L = -70mV$. The leak current has no gating variables.

The D-type potassium current is derived from Golomb et al., 2007 (7), and has a maximal conductance of $\bar{g}_D = 5\, mS/cm^2$. The D-current has three activation gates ($m$) and one inactivation gate ($h$) with the following kinetics:

$$m_\infty(V) = \frac{1}{1+\exp(-(V+50)/20)}$$
$$h_\infty(V) = \frac{1}{1+\exp((V+70)/6)}$$
$$\tau_m = 2$$
$$\tau_h = 150$$

The model of the A-type potassium current is derived from Traub et al., 2003 (8). The maximal conductance of the A-current is $\bar{g}_A = 3\, mS/cm^2$. The A-current has four activation gates ($m$) and one inactivation gate ($h$) with kinetics formulated as:

$$m_\infty = \frac{1}{1+\exp\left[-(V+60)/8.5\right]}$$
$$\tau_m = 0.185 + \frac{0.5}{\exp\left[(V+35.8)/19.7\right] + \exp\left[-(V+79.7)/12.7\right]}$$

$$h_\infty = \frac{1}{1+\exp\left[(V+78)/6\right]}$$

$$\tau_h = \begin{cases} \frac{0.5}{\exp[(V+46)/5]+\exp[-(V+238)/37.5]} & \text{if } V < -63mV \\ 9.5 & \text{if } V \geq -63mV \end{cases}$$

The model of the SOM cell h-current comes from Kramer et al., 2008 (9). The maximal h-current conductance is $\bar{g}_h = 2\, mS/cm^2$, and the h-current reversal potential is $E_h = -35mV$. The h-current has one activation gate ($r$) and no inactivation gate. The h-current activation variable steady state function and time constant of decay are given by:

$$r_\infty(V) = \frac{1}{1+\exp((V+75)/5.5}$$

$$\tau_r = \frac{1}{\exp(-14.6-0.086V)+\exp(-1.87+0.07V)}$$

The background excitation to each SOM cell was provided by giving an applied current of $I_{app} = 10\mu A/cm^2$. This value of Iapp put the SOM cell in its alpha-frequency stuttering mode at baseline (in the absence of any connections with other cells).

***VIP Cells***

Our VIP cell is modeled as the irregularly spiking (stuttering) type of VIP cell (10, 11). These cells are thought to have a D-type potassium current (11), an M-type potassium current (12), and a T-type calcium current (13). The voltage change of the VIP cells was modeled by the equation:

$$c_m \frac{dV}{dt} = -I_{Na} - I_K - I_L - I_D - I_M - I_T - I_{som_{vip}} - I_{pv_{vip}} - I_{e_{vip}} - I_{elec} + I_{app} \tag{8}$$

The sodium current ($I_{Na}$), the potassium current ($I_K$) and the D-type potassium current ($I_D$) are from Golomb et al., 2007 (7). The formulations of these currents are the same as the ones used for the SOM cell. The sodium current has a maximal conductance of $\bar{g}_{Na} = 112.5\, mS/cm^2$ and a reversal potential of $E_{Na} = 50mV$. The potassium current has a maximal conductance of $\bar{g}_K = 225\, mS/cm^2$. The potassium reversal potential is $E_K = -90mV$. The maximal conductance of the D-current is $\bar{g}_D = 3\, mS/cm^2$. The leak current maximal conductance is $\bar{g}_L = 0.25\, mS/cm^2$ and the leak current reversal potential is $E_L = -70mV$. The leak current has no gating variables. The background excitation is set to $I_{app} = 10\mu A/cm^2$.

The model of the M-current comes from Mainen et al., 1996 (4). The formulation of this current is the same as for the excitatory cell. The maximal conductance for the M-current ($\bar{g}_M$) is 1 mS/cm$^2$.

The dynamics of the T-type calcium current are governed by a constant field equation as in Huguenard and McCormick, 1992 (14).

$$I_T = Pz^2m^2h \cdot \frac{0.001F^2V}{RT} \cdot \frac{[Ca]_i - [Ca]_o \cdot \exp[-z0.001FV/(RT)]}{1.0 - \exp[-z0.001FV/(RT)]} \tag{9}$$

where $P = 0.02\, cm^3/s$ is the maximal permeability of calcium across the membrane, and $z = 2$ is the valence of calcium. The value of P is lower than that used in McCormick and Hueuenard, 1992 (15) but higher than that used in Wallenstein, 1994 (16). Faraday's constant is $F = 96489C/mole$. The temperature in Kelvin is $310K$, which is equivalent to a body temperature of $37\,^\circ C$. The gas constant is $R = 8.31\, Joules/(mole \cdot K)$. The internal calcium concentration ($[Ca]_i$) is $10\, nM$ and the external calcium concentration ($[Ca]_o$) is $3\,mM$.

The steady state activation curves and time constants of decay for the activation ($m$) and inactivation ($h$) gates are formulated similar to those in Huguenard and McCormick, 1992 (14):

$$m_\infty(V) = \frac{1}{1+\exp(-(V+57)/6.2}$$
$$h_\infty(V) = \frac{1}{1+\exp((V+81)/4}$$
$$\tau_m = \frac{1}{\exp(\frac{V+132}{-16.7})+\exp(\frac{V+16.8}{18.2})} + 0.612$$
$$\tau_h = \exp((V+467)/66.6), \qquad \text{if} V < -81mV$$
$$\tau_h = \exp(-(V+22)/10.5)+28, \qquad \text{if} V \geq -81mV$$

Synaptic currents ($I_{som_{vip}}$, $I_{pv_{vip}}$, $I_{e_{vip}}$) and electrical connections between VIP cells ($I_{elec}$) are discussed below.

***Parameter changes to simulate neuronal dysfunction (lowered neuronal excitability)***

We model excitatory deficiency in VIP by decreasing the VIP cell maximal sodium conductance to 80% of its original value. When decreased, $\bar{g}_{Na} = 90\,mS/cm^2$, which is low enough to decrease VIP excitability but not low enough to eliminate all VIP spiking during network activity. Our reason for choosing to decrease the activity of the sodium current ($I_{Na}$) rather than $I_{app}$ was to account for findings that classic Rett syndrome could be associated with mutations in SCN1A, which codes for a subunit in the voltage-gated sodium channel (17). We note that loss of function variants in the SCN1A gene are usually associated with Dravet syndrome (also known as severe myoclonic epilepsy in infancy), another severe neurodevelopmental disorder (18). Mouse models with SCN1A impairment have shown deficits in the voltage-gated sodium channels of irregularly spiking VIP cells (12), the type of VIP cell modeled in this study. Thus, we choose to model impairment of VIP cell excitability as a deficit in the sodium channel conductance.

For investigating the effect of lowered activity of other types of network neurons (SOM, PV or PYR) to PAC, we decreased the maximal sodium conductance for each particular neuron type to 80% of its original value while leaving the sodium conductances of all other neurons in the network at their original values.

***Synaptic connections***

We modeled two types of ionic synaptic currents ($I_{syn}$): AMPA currents ($I_{AMPA}$) from PYR and GABAa currents ($I_{GABAa}$) from all three interneuron subtypes. Both synaptic currents are derived or modified from Olufsen et al., 2003 (3) and are formulated as:

$$I_{syn} = \bar{g}s(V_{pre})(V - E_{ion}) \tag{10}$$

Each synaptic current has one activation gate (s), which depends on the voltage of the pre-synaptic neuron ($V_{pre}$). $E_{ion}$ is the synaptic reversal potential, and the maximal synaptic conductance is denoted by $\bar{g}$. The GABAa current is modeled by:

$$I_{GABAa} = \frac{\bar{g}_i}{N} s(V - E_i)$$

The maximal GABAa conductance ($\bar{g}_i$) of a neuron is normalized by the number ($N$) of presynaptic neurons of a specific interneuron type projecting to that neuron. The value of $\bar{g}_i$ depends on the pre- and post-synaptic neurons. The value of $\bar{g}_i$ in units of $mS/cm^2$ for each pre- and post-synaptic pair is: $\bar{g}_{VIP-to-PV} = 0.1$, $\bar{g}_{PV-to-VIP} = 0.01$, $\bar{g}_{VIP-to-SOM} = 0.08$, $\bar{g}_{SOM-to-VIP} = 0.5$, $\bar{g}_{SOM-to-PV} = 0.1$, $\bar{g}_{PV-to-SOM} = 0.1$, $\bar{g}_{PV-to-PV} = 0.2$, $\bar{g}_{PV-to-PYR} = 0.4$, $\bar{g}_{SOM-to-PYR} = 0.2$, and $\bar{g}_{VIP-to-PYR} = 0.01$. The reversal potential for the GABAa current ($E_i$) is set to $-80$ mV for all neurons.

The activation gate ($s$) of each GABAa current has kinetics described by:

$$\frac{ds}{dt} = \bar{g}(V_{pre})(1 - s) - s/\tau_i \tag{11}$$

For most GABAa synaptic connections $\bar{g}(V_{pre})$ is formulated as:

$$\bar{g}(V_{pre}) = 2(1 + tanh(\frac{V_{pre}}{4}))$$

The exceptions occur for SOM-to-PV connections, where $\bar{g}(V_{pre})$ is

$$\bar{g}(V_{pre}) = 2.5(1 + tanh(\frac{V_{pre}}{0.1}))$$

and for SOM-to-VIP and SOM-to-PYR connections, where $\bar{g}(V_{pre})$ is:

$$\bar{g}(V_{pre}) = 5(1 + tanh(\frac{V_{pre}}{0.1}))$$

The time constant of decay ($\tau_i$) of the activation gate ($s$) depends on the type of pre- and post-synaptic neurons. The value of $\tau_i$ in units of milliseconds for each pre- and post-synaptic neuron is: $\tau_{PV-to-VIP} = 18$, $\tau_{PV-to-PYR} = 8$, $\tau_{PV-to-PV} = 8$, $\tau_{PV-to-SOM} = 18$, $\tau_{VIP-to-SOM} = 40$, $\tau_{VIP-to-PV} = 6$, $\tau_{VIP-to-PYR} = 40$, $\tau_{SOM-to-PV} = 40$, $\tau_{SOM-to-VIP} = 20$, $\tau_{SOM-to-PYR} = 20$. Values of $\tau_i$ were taken from the literature when found (19–21).

The AMPA current is modeled by:

$$I_{AMPA} = \frac{\bar{g}_e}{N} s(V - E_e)$$

The maximal AMPA conductance ($\bar{g}_e$) of a neuron is normalized by the number ($N$) of presynaptic excitatory neurons projecting to that neuron. The value of $\bar{g}_e$ depends on the type of post-synaptic neuron. The value of $\bar{g}_e$ in units of $mS/cm^2$ for each pre- and post-synaptic pair is: $\bar{g}_{PYR-to-PV} = 0.4$, $\bar{g}_{PYR-to-VIP} = 0.3$, $\bar{g}_{PYR-to-SOM} = 0.1$. Additionally, we included extremely weak synaptic currents between PYR cells ($\bar{g}_{PYR-to-PYR} = 0.001$) in order to generate an approximation of the LFP, which was modeled as the sum of all PYR-to-PYR AMPA currents in the network. The reversal potential for the AMPA current ($E_e$) is set to 0 mV for all neurons.

The activation gate ($s$) of each AMPA current has kinetics described by:

$$\frac{ds}{dt} = \bar{g}(V_{pre})(1 - s) - s/\tau_e \tag{12}$$

$$\bar{g}(V_{pre}) = 5(1 + tanh(\frac{V_{pre}}{4}))$$

The value of $\tau_e$ was set to 2 ms for all AMPA synapses.

Electrical connections ($I_{elec}$): Electrical connections (gap junctions) were present between all interneurons of the same type (21, 22). The gap junction current from interneuron j to interneuron i was modeled as:

$$I_{elec} = \bar{g}_{elec}(V_i - V_j)$$

The maximal gap junction conductance ($\bar{g}_{elec}$) was 0.005 $mS/cm^2$ between all neurons of the same type.

***Networks***

Our networks consisted of 110 neurons: 90 PYR, 11 PV+IN, 6 SOM+IN and 3 VIP+IN. These numbers were chosen based on an approximate proportion of each cell type found in cortex (23). All-to-all connections were used from SOM, PV and VIP to excitatory cells (PYR). Each PV+IN received AMPA input from 50 randomly chosen PYR. SOM+IN and VIP+INs received input from distinct sets of PYR (21). Each VIP+IN received input from 45 PYR that did not send projections to SOM+INs, and each SOM+IN received input from 6 PYR randomly selected from the 45 PYR that did not project to VIP+INs. PV+INs were connected all-to-all with each other, excluding autapses. Similarly, PYR were connected all to-all with each other (excluding autapses) with very weak connections used to calculate the model LFP. SOM+INs and VIP+INs had no synaptic connections between members of their own subtype. Electrical connections among interneurons of the same type were all-to-all, and synaptic connections between all different subtypes of interneurons were all-to-all.

### Tables S1–S2

**Table S1. Age- and power-adjusted group comparisons**

| Outcome | Region | Age-adjusted group effect | Power-adjusted group effect |
|---|---|---|---|
| **PAC strength, theta-gamma** | Whole brain | **0.67 [0.20, 1.14] p=.006; q=.022** | **0.61 [0.11, 1.12] p=.018; q=.030** |
| | Anterior | 0.42 [−0.16, 0.99] p=.154; q=.190 | 0.36 [−0.27, 0.99] p=.258; q=.295 |
| | Posterior | **1.05 [0.39, 1.72] p=.002; q=.013** | **0.85 [0.14, 1.56] p=.020; q=.030** |
| | Midline | **0.38 [0.09, 0.66] p=.010; q=.025** | **0.36 [0.07, 0.65] p=.017; q=.030** |
| **PAC strength, alpha-gamma** | Whole brain | **1.00 [0.40, 1.60] p=.001; q=.011** | **0.91 [0.21, 1.60] p=.011; q=.025** |
| | Anterior | **0.69 [0.11, 1.27] p=.021; q=.030** | 0.63 [−0.01, 1.27] p=.053; q=.071 |
| | Posterior | **1.80 [0.92, 2.67] p<.001; q=.002** | **1.43 [0.35, 2.51] p=.010; q=.025** |
| | Midline | 0.04 [−0.78, 0.87] p=.915; q=.915 | 0.32 [−0.63, 1.28] p=.501; q=.535 |

Values are adjusted RTT-minus-TD coefficients [95% CI], with positive values indicating higher PAC strength in RTT, followed by nominal *p*-values and Benjamini–Hochberg-adjusted *q*-values. Estimates were obtained using ordinary linear regression. Age-adjusted models included diagnostic group and mean-centered age. Power-adjusted models included diagnostic group and same-region log10-transformed theta and gamma PSD for theta–gamma PAC, or alpha and gamma PSD for alpha–gamma PAC. FDR correction was applied jointly across the 16 adjusted group comparisons: eight age-adjusted and eight power-adjusted group effects. Bold values indicate $q < .05$.

**Table S2. Medication use among RTT participants**

| Theta-gamma PAC strength | Alpha-gamma PAC strength | Medications |
|---|---|---|
| 3.51 | 6.62 | Hydroxyzine |
| 3.32 | 5.37 | Polyethylene glycol; Risperidone; Trazodone |
| 5.21 | 3.19 | Budesonide; Clonazepam; Clonidine; Diazepam (PRN); Lamotrigine; Levalbuterol; Levetiracetam; Melatonin; Montelukast; Multivitamin; Polyethylene glycol |
| 4.68 | 3.54 | Cholecalciferol; Ferrous sulfate |
| 3.24 | 4.41 | Escitalopram |
| 1.24 | 5.57 | Acetylcysteine; Baclofen; Cannabidiol; Carbamide peroxide otic solution; Cholecalciferol; Clonidine; Diazepam (PRN); Esomeprazole; Famotidine; Gabapentin; Hydrocortisone ointment; Hydroxyzine; Melatonin; Norethindrone; Polyethylene glycol; Prucalopride; Risperidone; Topiramate; Trofinetide |
| 3.08 | 3.48 | Melatonin |
| 2.10 | 4.03 | Diazepam; Fluticasone nasal spray; Gabapentin; Polyethylene glycol |
| 2.03 | 3.77 | Clonidine; Fluoxetine |
| 2.30 | 2.72 | Clonidine; Gabapentin; Levocarnitine; Mirtazapine; Trazodone |
| 2.16 | 2.83 | Buspirone; Cetirizine; Clonidine; Diphenhydramine (PRN); Fluticasone nasal spray; Melatonin; Multivitamin; Probiotic; Trazodone; Trofinetide |
| 1.22 | 2.62 | Cholecalciferol; Diazepam (PRN) |
| 1.60 | 1.65 | Cetirizine; Magnesium; Melatonin; Oral nutritional supplement with fiber; Riboflavin; Ubiquinone |
| 1.09 | 2.02 | Cholecalciferol; Cyproheptadine; Trofinetide |
| 1.16 | 1.94 | Lamotrigine |
| 1.05 | 1.81 | Albuterol; Cyproheptadine; Fluticasone inhaler; Glycerin suppository; Magnesium hydroxide; Senna; Topiramate |
| 1.06 | 1.52 | Butyrate supplement; Levetiracetam; Multivitamin; Omega-3 fatty acids; Probiotic |
| 1.18 | 1.35 | Cholecalciferol; Levetiracetam; Magnesium; Omega-3 fatty acids; Polyethylene glycol |
| 1.19 | 1.04 | Cannabidiol; Melatonin (PRN); Oxcarbazepine |
| 1.02 | 1.11 | Glycopyrrolate; Lacosamide; Levocarnitine; Polyethylene glycol; Valproic acid |
| 0.75 | 1.35 | Clonidine; Famotidine; Probiotic |
| 1.02 | 1.05 | Albuterol (PRN); Atropine ophthalmic; Bisacodyl; Buspirone; Docusate; Melatonin; Mirtazapine; Propranolol |
| 0.86 | 1.07 | Buspirone; Cannabidiol; Cannabis; Clonazepam (PRN); Clonidine; Ethinyl estradiol/norethindrone; Lorazepam (PRN); Melatonin; Midazolam (PRN); Phenytoin; Prucalopride |
| 0.68 | 1.13 | Clonidine; Guanfacine; Mirtazapine; Trazodone |
| 0.79 | 0.83 | Cetirizine; Creatine; Melatonin; Polyethylene glycol |
| 0.66 | 0.94 | Cholecalciferol; Cyproheptadine; Ethinyl estradiol/levonorgestrel; Glycopyrrolate; Polyethylene glycol; Sertraline; Topiramate |
| 0.76 | 0.82 | Levetiracetam |
| 0.60 | 0.84 | Clonazepam; Polyethylene glycol; Potassium citrate/citric acid; Zonisamide |
| 0.50 | 0.89 | Calcium/cholecalciferol; Cholecalciferol; Diazepam (PRN); Ethinyl estradiol/norethindrone; Lamotrigine; Levocarnitine; Multivitamin; Omega-3 fatty acids; Polyethylene glycol |

**Table S2. Medication use among RTT participants**

| Theta-gamma PAC strength | Alpha-gamma PAC strength | Medications |
|---|---|---|
| 0.70 | 0.68 | Clonazepam; Clonidine; Diazepam (PRN); Glycopyrrolate; Levetiracetam; Lorazepam (PRN); Sertraline |
| 0.54 | 0.78 | Polyethylene glycol; Trofinetide |
| 0.49 | 0.81 | Cholecalciferol; Clonidine; Ferrous sulfate; Lansoprazole; Magnesium; Polyethylene glycol; Probiotic |
| 0.26 | 0.99 | Lamotrigine; Levetiracetam; Polyethylene glycol |
| 0.55 | 0.49 | Loperamide; Trofinetide |
| 0.43 | 0.28 | Ammonium lactate lotion; Bethanechol; Bisacodyl; Diazepam (PRN); Oxygen |
| 0.44 | 0.17 | Esomeprazole; Multivitamin; Ranitidine |

Includes participants with one or more reported medications (we were unable to obtain medication records from two participants). Rows are ordered from highest to lowest mean PAC strength, using whole-brain theta-gamma and whole-brain alpha gamma MInorm values. Medication names listed alphabetically.

### Figures S1–S3

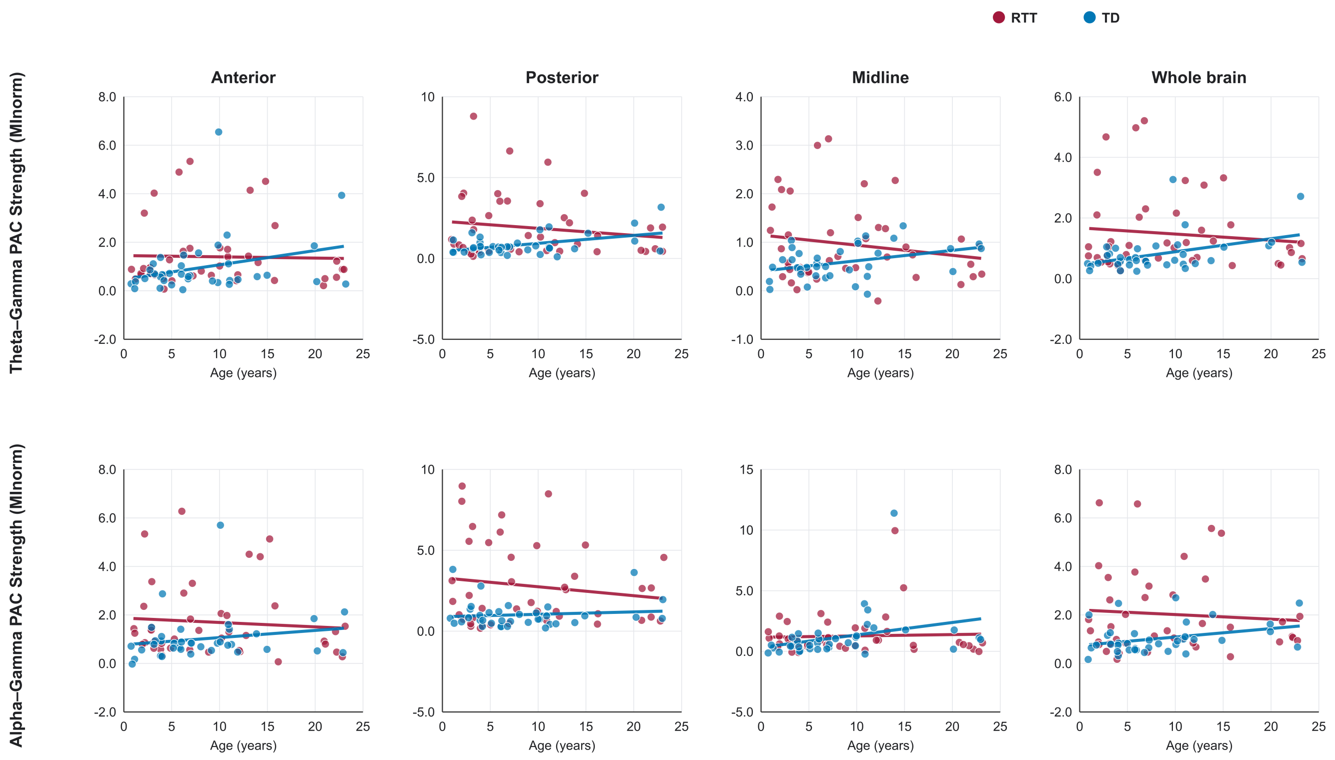


**Figure S1. Relationships between age and PAC strength in RTT and TD.** Each point represents one participant's normalized PAC strength (MI*norm*) for theta–gamma PAC (top) or alpha–gamma PAC (bottom) in the anterior, posterior, midline, and whole-brain regions. RTT participants (n = 42) are shown in red and TD participants (n = 37) in blue. Lines show within-group linear regression fits over each group's observed age range.

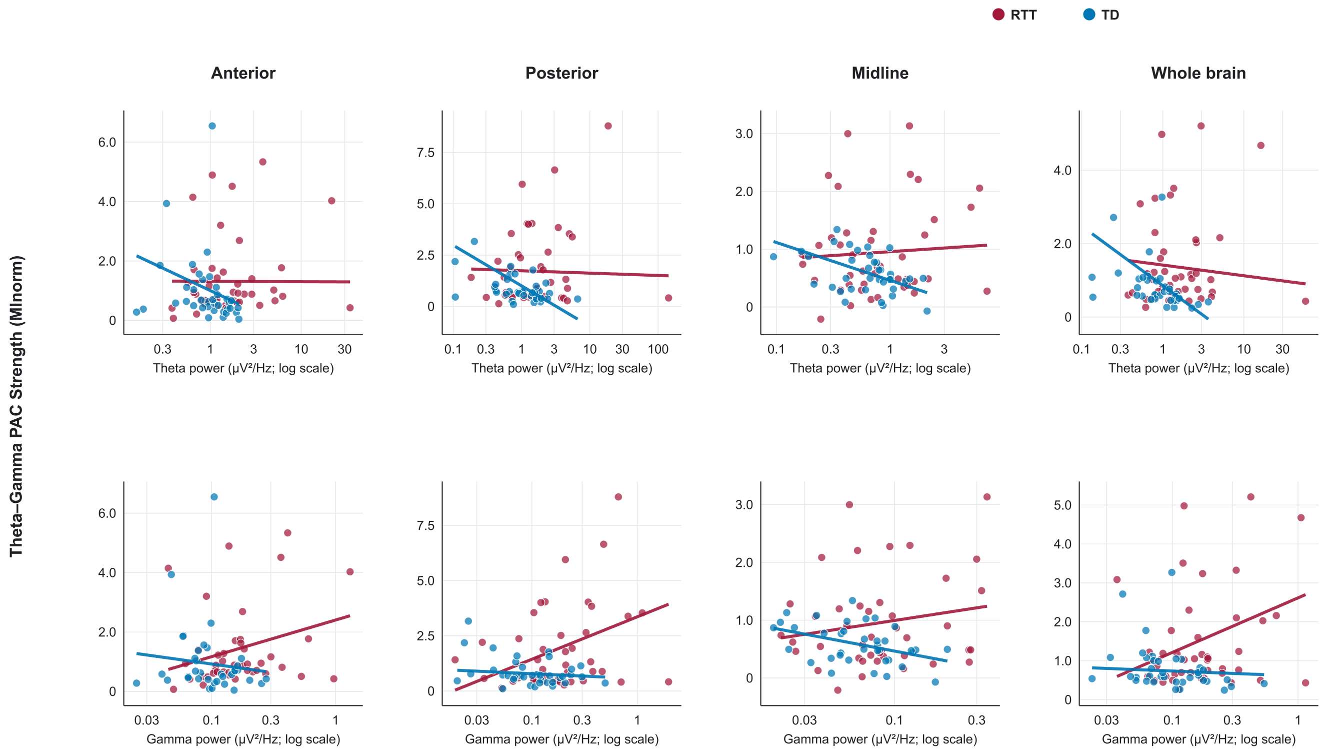


**Figure S2. Relationships between spectral power and theta–gamma PAC strength in RTT and TD.** Each point represents one participant's normalized theta–gamma PAC strength (MI*norm*) in the anterior, posterior, midline, or whole-brain region. Theta power is shown in the top row and gamma power in the bottom row. Band-averaged PSD (μV²/Hz) is displayed on a logarithmic scale. RTT participants (n = 42) are shown in red and TD participants (n = 37) in blue. Lines show fitted group-specific relationships from models containing both regional theta and gamma power; the other power term was held at its sample mean, and lines are limited to each group's observed power range.

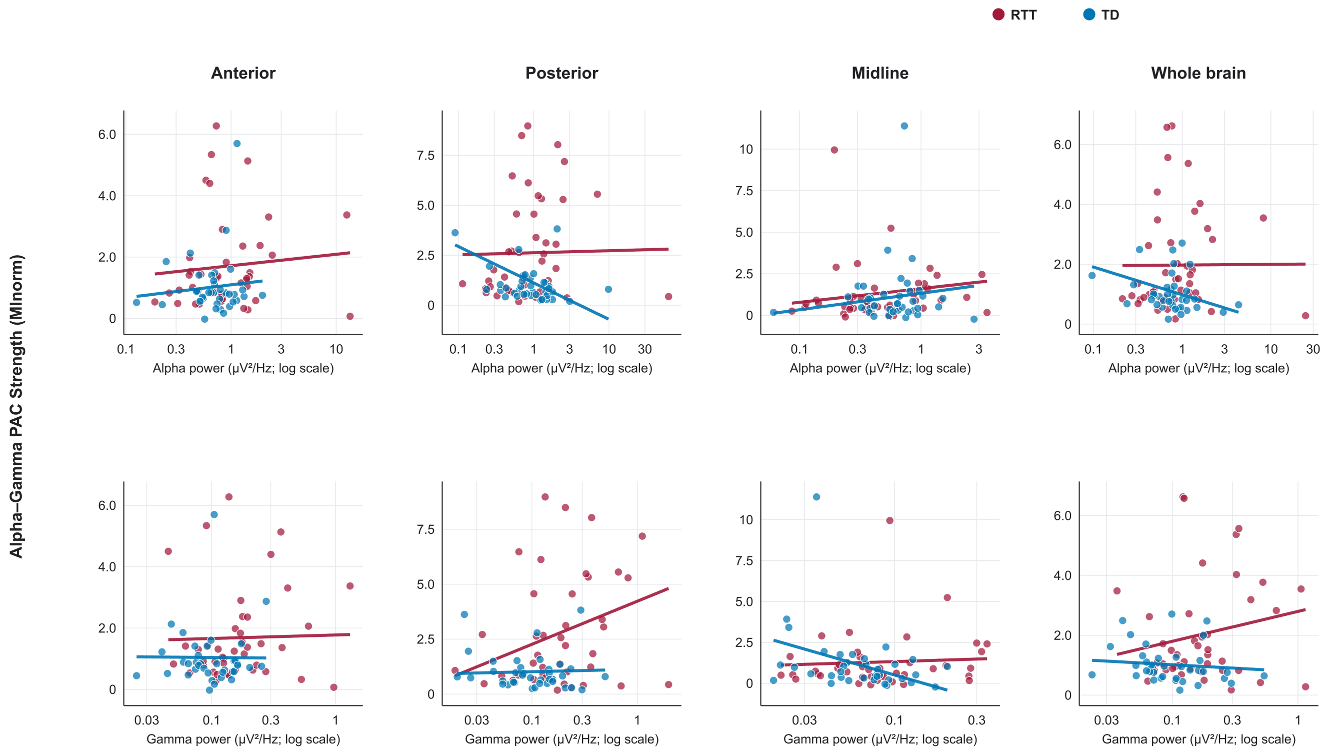


**Figure S3. Relationships between spectral power and alpha–gamma PAC strength in RTT and TD.** Each point represents one participant's normalized alpha–gamma PAC strength (MI*norm*) in the anterior, posterior, midline, or whole-brain region. Alpha power is shown in the top row and gamma power in the bottom row. Band-averaged PSD (µV²/Hz) is displayed on a logarithmic scale. RTT participants (n = 42) are shown in red and TD participants (n = 37) in blue. Lines show fitted group-specific relationships from models containing both regional alpha and gamma power; the other power term was held at its sample mean, and lines are limited to each group's observed power range.
